# Sleeping under the waves: a longitudinal study across the contagion peaks of COVID-19 pandemic in Italy

**DOI:** 10.1101/2021.01.17.21249947

**Authors:** Federico Salfi, Aurora D’Atri, Daniela Tempesta, Michele Ferrara

## Abstract

After the March–April 2020 COVID-19 outbreak, a second contagion wave afflicted Europe in autumn. This study aimed to evaluate sleep health/patterns of Italians during this further challenging situation.

A total of 2013 Italians longitudinally participated in a web-based survey during the two contagion peaks of the COVID-19 outbreak. We investigated the risk factors for sleep disturbances during the second wave, and we compared sleep quality and psychological well-being between the two assessments (March–April and November–December 2020). Female gender, low education, evening chronotype, being at high-risk for COVID-19 infection, reporting negative social or economic impact, and evening smartphone overuse predicted a higher risk of poor sleep and insomnia symptoms during the second wave. Advanced age, living with high-risk subjects for COVID-19 infection, and having a relative/friend infected with COVID-19 before the prior two weeks were risk categories for poor sleep quality. Living with children, having contracted COVID-19 before the prior two weeks, being pessimistic on the vaccine, and working in healthcare were risk factors for insomnia symptoms. The follow-up assessment highlighted reduced insomnia symptoms and anxiety. Nevertheless, we showed reduced sleep duration, higher daytime dysfunction and sleep medication use, and advanced sleep phase, confirming the alarming prevalence of poor sleepers (∼60%) and severe depression (∼20%) in a context of increased perceived stress.

This study demonstrated a persistent impact of the COVID-19 pandemic on sleep and mental health. Large-scale interventions to counteract the chronicity and exacerbation of sleep and psychological disturbances are necessary, especially for the risk categories.

## Introduction

From December 2019, Coronavirus disease 2019 (COVID-19) began to spread worldwide rapidly. The Italian government reacted to the first contagion wave (March–April 2020) implementing a total lockdown involving home confinement and social distancing for the entire population, and the closure of most business activities. The lockdown started on 9 March and lasted until 3 May 2020. Many studies demonstrated a pervasive impact of the lockdown period during the first wave of COVID-19 outbreak on sleep and psychological health of the general population (Jahrami et al., 2020; Rakiumar et al., 2020). In autumn, a second contagion outbreak occurred in Italy, leading the government to adopt new restraining measures to control the virus propagation. A regional approach was adopted this time: restrictions of movement freedom, business and school activities have been imposed according to the local infection diffusion and the pressure on regional healthcare system. On 6 November 2020, Italian regions have been classified across three risk levels (yellow, orange, and red), periodically updated based on the COVID-19 related data monitoring. In the current study, we provide the first evaluation of sleep health of the general population during the second wave of COVID-19 emergency, identifying the at-risk categories for sleep disturbances during this further challenging period. Moreover, we longitudinally compared the outcome of the current period in a large Italian sample with the situation of the first pandemic wave, through a web-based survey administered the two weeks around the two contagions peaks.

## Methods

### Participants and Procedure

The present study is part of a larger research project aimed at understanding the consequences of COVID-19 outbreak on the Italian population (Salfi et al., 2020a, 2020b). A total of 8798 Italian citizens participated in a web-based survey during the first wave of COVID-19 (Test 1: 25 March–7 April 2020; Figure 1), corresponding to the two weeks centred on the contagion peak (the third and fourth week of lockdown). The survey evaluated sleep quality, insomnia symptoms, chronotype, depression symptoms, perceived stress, and anxiety using the following validated questionnaires: the Pittsburgh Sleep Quality Index (PSQI; Curcio et al., 2013), the Insomnia Severity Index (ISI; Castronovo et al., 2016), the reduced form of the Morningness-Eveningness Questionnaire (MEQr; Natale et al., 2006), the Beck Depression Inventory-second edition (BDI-II; Ghisi et al., 2006), the 10-item Perceived Stress Scale (PSS-10; Mondo et al., 2019), and the state-anxiety subscale of the State-Trait Anxiety Inventory (STAI-X1; Spielberg et al., 1970).

**Figure 1.**
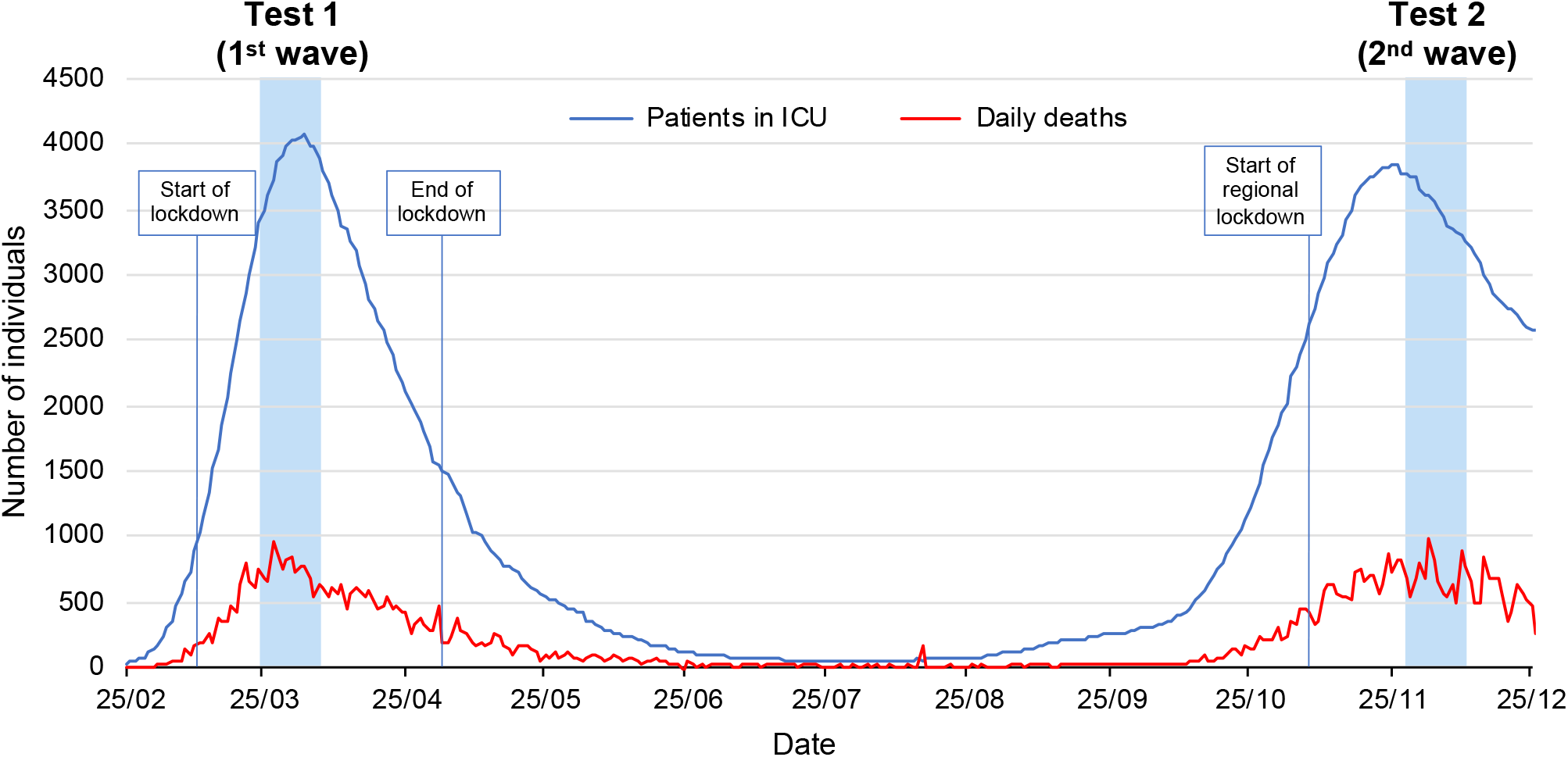
Italian national trend of daily deaths and patients in Intensive Care Unit (ICU) due to COVID-19 infection across the pandemic period (Protezione civile, 2020). The two assessment periods (Test 1: 25 March–7 April 2020, Test 2: 28 November–11 December 2020) are marked by the light blue areas.

The Test 1 respondents were invited by email to participate in a follow-up assessment on 28 November 2020, corresponding to the contagion peak of the second wave of COVID-19 outbreak. A total of 2013 individuals participated in the second measurement in a two-week time window (Test 2: 28 November–11 December 2020; Figure 1). The follow-up measurement comprised the same questionnaires as Test 1. Additionally, we collected the following sociodemographic and COVID-19-related information as continuous or categorical variables: age, gender, education, occupation, geographic region used to derive the restraining measures in force (yellow, orange, or red zone), living with children, being high-risk subject for COVID-19 infection, living with high-risk subjects for COVID-19 infection, COVID-19 infection, forced quarantine, infection or death of a relative/close friend due to COVID-19, perspective on vaccination, negative economic and social impact of the current situation, and mean exposure (min) to smartphone, PC/tablet, TV, and e-reader in the two hours before falling asleep during the last two weeks. The available choices for each categorical variable along with the sociodemographic composition of the follow-up sample, and the COVID-related responses are reported in Table 1. The compilation of the last three questionnaires (BDI-II, PSS-10, STAI-X1) was optional to ensure reliable unforced responses. A total of 1847, 1790, and 1784 participants compiled the BDI-II, the PSS-10, and the STAI-X1, respectively, during Time 1 and Time 2. Online informed consent was obtained from participants. The study has been approved by the Internal Review Board of the University of L’Aquila (protocol n. 43066) and has been carried out according to the principles established by the Declaration of Helsinki.

**Table 1.** Sociodemographic composition and COVID-related responses of the Test 2 sample and results (B, Odds ratio, 95% confidence interval, P) of the logistic regression analyses on PSQI (>5) and ISI (>14) scores.

| Predictor | N (%)<br>[*Mean (SD)] | Poor sleep<br>(PSQI >5) |  |  | Moderate/Severe Insomnia<br>(ISI >14) |  |  |
| --- | --- | --- | --- | --- | --- | --- | --- |
|  |  | B | OR (95% CI) | P | B | OR (95% CI) | P |
| **Intercept** |  | -1.99 | 0.14 (0.06–0.32) | **< 0.001** | -4.36 | 0.01 (0.002–0.07) | **< 0.001** |
| **Age** | *34.84<br>(12.37) | 0.02 | 1.02 (1.01–1.03) | **< 0.01** | -0.01 | 0.99 (0.97–1.01) | 0.35 |
| **Gender** |  |  |  |  |  |  |  |
| Female | 1648 (81.87) | Reference |  |  | Reference |  |  |
| Male | 365 (18.13) | -0.39 | 0.68 (0.52–0.87) | **< 0.01** | -0.45 | 0.64 (0.41–0.998) | **< 0.05** |
| **Education** |  |  |  |  |  |  |  |
| Middle/High school | 586 (29.11) | Reference |  |  | Reference |  |  |
| Graduated | 1163 (57.77) | -0.22 | 0.80 (0.64–1.01) | 0.06 | -0.47 | 0.62 (0.45–0.87) | **< 0.01** |
| Over graduated | 264 (13.12) | -0.33 | 0.72 (0.51–0.99) | **< 0.05** | -0.31 | 0.73 (0.44–1.21) | 0.22 |
| **Occupation** |  |  |  |  |  |  |  |
| Unemployed | 151 (7.50) | Reference |  |  | Reference |  |  |
| Student | 521 (25.88) | -0.20 | 0.82 (0.53–1.28) | 0.38 | 0.13 | 1.13 (0.60–2.13) | 0.70 |
| Healthcare worker | 159 (7.90) | -0.36 | 0.70 (0.42–1.18) | 0.18 | 0.86 | 2.36 (1.15–4.83) | **0.02** |
| Self-employed | 340 (16.89) | -0.33 | 0.72 (0.46–1.13) | 0.15 | 0.31 | 1.36 (0.71–2.60) | 0.35 |
| Employed | 772 (38.35) | -0.16 | 0.86 (0.56–1.30) | 0.46 | 0.43 | 1.53 (0.84–2.78) | 0.16 |
| Retired | 70 (3.48) | -0.26 | 0.77 (0.36–1.66) | 0.51 | 0.86 | 2.36 (0.76–7.37) | 0.14 |
| **Restraining measures** |  |  |  |  |  |  |  |
| Red zone | 1046 (51.96) | Reference |  |  | Reference |  |  |
| Orange zone | 451 (22.40) | -0.07 | 0.93 (0.73–1.20) | 0.58 | 0.11 | 1.12 (0.77–1.62) | 0.55 |
| Yellow zone | 516 (25.63) | 0.05 | 1.06 (0.83–1.34) | 0.66 | -0.08 | 0.92 (0.64–1.33) | 0.67 |
| **Chronotype** |  |  |  |  |  |  |  |
| Neither-type | 1289 (64.03) | Reference |  |  | Reference |  |  |
| Morning-type | 487 (24.19) | -0.44 | 0.65 (0.51–0.82) | **< 0.001** | -0.48 | 0.62 (0.41–0.94) | **0.03** |
| Evening-type | 237 (11.77) | 0.42 | 1.53 (1.10–2.13) | **0.01** | 0.71 | 2.03 (1.36–3.03) | **< 0.001** |
| **Living with children** |  |  |  |  |  |  |  |
| No | 1566 (77.79) | Reference |  |  | Reference |  |  |
| Yes | 447 (22.21) | 0.12 | 1.13 (0.89–1.45) | 0.32 | 0.41 | 1.50 (1.06–2.13) | **0.02** |
| **Being high-risk subject for COVID-19 infection** |  |  |  |  |  |  |  |
| No | 1812 (90.02) | Reference |  |  | Reference |  |  |
| Yes | 201 (9.99) | 0.49 | 1.64 (1.13–2.38) | **< 0.01** | 0.59 | 1.80 (1.16–2.81) | **< 0.01** |
| **Living with high-risk subject for COVID-19 infection** |  |  |  |  |  |  |  |
| No | 1493 (74.17) | Reference |  |  | Reference |  |  |
| Yes | 520 (25.83) | 0.31 | 1.36 (1.07–1.71) | **0.01** | 0.01 | 1.01 (0.72–1.41) | 0.96 |
| **COVID-19 infection** |  |  |  |  |  |  |  |
| No | 1887 (93.74) | Reference |  |  | Reference |  |  |
| Yes (last two weeks) | 34 (1.69) | 0.92 | 2.50 (0.87–7.17) | 0.09 | 0.84 | 2.32 (0.78–6.86) | 0.13 |
| Yes, (before last two weeks) | 92 (4.57) | 0.18 | 1.20 (0.71–2.02) | 0.50 | 0.66 | 1.94 (1.02–3.70) | **0.04** |
| **Forced quarantine** |  |  |  |  |  |  |  |
| No | 1513 (75.16) | Reference |  |  | Reference |  |  |
| Yes (last two weeks) | 128 (6.36) | 0.12 | 1.13 (0.70–1.81) | 0.62 | 0.01 | 1.01 (0.51–2.01) | 0.98 |
| Yes, (before last two weeks) | 372 (18.48) | -0.02 | 0.98 (0.74–1.30) | 0.89 | 0.05 | 1.05 (0.69–1.58) | 0.83 |

|  |  | Poor sleep<br>(PSQI >5) |  |  | Moderate/Severe Insomnia<br>(ISI >14) |  |  |
| --- | --- | --- | --- | --- | --- | --- | --- |
| Predictor | N (%)<br>[*Mean (SD)] | B | OR (95% CI) | P | B | OR (95% CI) | P |
| COVID-19 infection of a relative/close friend |  |  |  |  |  |  |  |
| No | 823 (40.88) | Reference |  |  | Reference |  |  |
| Yes (last two weeks) | 541 (26.88) | 0.17 | 1.19 (0.92–1.52) | 0.19 | -0.02 | 0.99 (0.68–1.44) | 0.94 |
| Yes, (before last two weeks) | 649 (32.24) | 0.31 | 1.36 (1.06–1.74) | **0.01** | -0.10 | 0.90 (0.62–1.30) | 0.59 |
| Death of a relative/close friend due to COVID-19 |  |  |  |  |  |  |  |
| No | 1832 (91.01) | Reference |  |  | Reference |  |  |
| Yes | 181 (8.99) | -0.04 | 0.96 (0.67–1.37) | 0.82 | 0.06 | 1.06 (0.64–1.77) | 0.81 |
| Perspective on vaccination |  |  |  |  |  |  |  |
| Optimistic | 853 (42.38) | Reference |  |  | Reference |  |  |
| Undecided | 887 (44.06) | 0.19 | 1.21 (0.98–1.49) | 0.08 | 0.29 | 1.33 (0.95–1.86) | 0.09 |
| Pessimistic | 273 (13.56) | 0.20 | 1.22 (0.89–1.67) | 0.23 | 0.61 | 1.84 (1.20–2.81) | **< 0.01** |
| Economic impact |  |  |  |  |  |  |  |
| None | 1055 (52.41) | Reference |  |  | Reference |  |  |
| Negative | 712 (35.37) | 0.47 | 1.60 (1.27–2.01) | **< 0.001** | 0.57 | 1.78 (1.28–2.46) | **< 0.001** |
| Positive | 246 (12.22) | -0.08 | 0.92 (0.68–1.26) | 0.61 | -0.17 | 0.85 (0.49–1.47) | 0.56 |
| Negative social impact |  |  |  |  |  |  |  |
| None | 74 (3.68) | Reference |  |  | Reference |  |  |
| A little | 760 (37.76) | 0.79 | 2.21 (1.28–3.80) | **< 0.01** | 0.85 | 2.35 (0.55–10.02) | 0.25 |
| A lot | 1179 (58.57) | 1.46 | 4.29 (2.50–7.36) | **< 0.001** | 1.59 | 4.93 (1.18–20.65) | **0.03** |
| Evening electronic device usage (min) |  |  |  |  |  |  |  |
| Smartphone | *55.25 (37.96) | 0.009 | 1.009 (1.006–1.012) | **< 0.001** | 0.009 | 1.009 (1.005–1.013) | **< 0.001** |
| PC and tablet | *37.02 (44) | 0.002 | 1.002 (1.00–1.005) | 0.11 | 0.001 | 1.001 (0.998–1.004) | 0.57 |
| Television | *50.11 (45.96) | -0.001 | 0.999 (0.996–1.001) | 0.27 | 0.001 | 0.999 (0.996–1.003) | 0.70 |
| E-reader | *6.08 (19.53) | 0.003 | 1.003 (0.997–1.008) | 0.32 | 0.002 | 1.002 (0.996–1.009) | 0.51 |

### Statistical analysis

Prevalence of poor sleepers, moderate/severe insomnia symptoms, and severe depression condition were computed according to the conventional cut-off scores: PSQI >5, ISI >14, BDI-II >28, respectively. MEQr scores were used to assign the respondents to three chronotype groups (Morning-type: 19–25; Neither-type: 11–18; Evening-type: 4–10).

In order to provide a comprehensive overlook on sociodemographic and COVID-related factors influencing the risk for poor sleep quality and moderate/severe insomnia during the second pandemic wave, we performed binomial logistic regressions on PSQI (>5) and ISI (>14) scores including the available sociodemographic and COVID-19-related variables as predictors.

The investigation on possible changes in sleep quality, insomnia symptoms, chronotype, depression, perceived stress, and anxiety among the two infection waves has been performed contrasting the questionnaire scores (PSQI, ISI, MEQr, BDI-II, PSS-10, STAI-X1, respectively) at the two time-points by paired t-tests. The analysis has been replicated for specific items (bedtime and wake-up time) and each component of the PSQI (Subjective sleep quality, Sleep latency, Sleep duration, Habitual sleep efficiency, Sleep disturbances, Sleep medications, Daytime dysfunction) to further detail the specific dimensions of sleep habits/quality possibly changed between the two assessments.

The prevalence of poor sleep quality, moderate/severe insomnia, and severe depression have been compared between the two time-points using McNemar test.

All tests were two-tailed and statistical significance was set to P<0.05. We excluded 153 respondents from the PSQI analyses, due to compilation errors.

## Results

### At-risk categories for sleep disturbances during the second wave

The results of the logistic regression models are reported in Table 1. Female gender, evening chronotype, being a COVID-19 high-risk subject, reporting negative social or economic impact of the current situation, and higher smartphone usage in the two hours before falling asleep predicted a higher risk of poor sleep and moderate/severe insomnia symptoms during the COVID-19 second wave. On the other hand, higher education level and morning chronotype emerged as protective factors against poor sleep quality and insomnia symptoms.

Advanced age, living with a high-risk subject for infection, and having a relative/close friend infected with COVID-19 were at-risk categories for poor sleep quality, while living with children, having contracted COVID-19 before the last two weeks, being pessimistic on the vaccination prospective, and being a healthcare worker were risk factors for developing moderate/severe insomnia symptoms.

### Sleep and psychological differences between the waves

Results of t-test comparisons between Test 1 and Test 2 are reported in Table 2. The sample went to bed and woke-up earlier at Test 2. Notwithstanding the lack of significant changes on PSQI total score, the analyses on PSQI components highlighted several differences between the two measurements. Subjective sleep quality, sleep latency, and sleep disturbances improved. On the contrary, respondents slept less, increased sleep medications use, and showed higher daytime dysfunction. Moreover, the participants showed reduced severity of insomnia symptoms, and increased MEQr scores, pointing to morning chronotype. Finally, depressive symptomatology remained stable, while perceived stress increased, and anxiety declined. The prevalence comparisons showed that the percentage of poor sleepers remained stable, insomniacs declined, and the individuals reporting severe depression symptoms remained unchanged over time.

**Table 2.** Descriptive statistics (mean and standard deviation) of questionnaire scores assessing sleep/chronobiological habits and quality (bedtime and wake-up time, PSQI total score and sub-components, ISI, MEQr), and psychological condition (BDI-II, PSS-10, STAI-X1) for Test 1 (25 March–7 April) and Test 2 (28 November–11 December), and the corresponding statistical comparisons (paired t-test: t, P). Prevalence (%) of sleep disturbances (poor sleep and moderate/severe insomnia) and severe depression at the two time points and the corresponding statistical comparisons (McNemar: χ^2^, P) are also reported.

|  | Test 1<br>(1<sup>st</sup> wave) | Test 2<br>(2<sup>nd</sup> wave) |  |  |
| --- | --- | --- | --- | --- |
| *Sleep/chronotype features* | Mean (SD) |  | t | P |
| Bedtime (hh:mm) | 00:14 (1:25) | 11:38 (1:17) | 21.64 | **< 0.001** |
| Wake-up time (hh:mm) | 8:44 (1:38) | 7:50 (1:22) | 28.15 | **< 0.001** |
| PSQI total score | 6.95 (3.67) | 6.90 (3.54) | 0.58 | 0.56 |
| Subjective sleep quality | 1.39 (0.78) | 1.34 (0.72) | 2.57 | **0.01** |
| Sleep latency | 1.43 (1.04) | 1.27 (1.02) | 6.63 | **< 0.001** |
| Sleep duration | 0.70 (0.81) | 0.79 (0.78) | -5.25 | **< 0.001** |
| Habitual sleep efficiency | 0.79 (1.00) | 0.81 (1.00) | -0.60 | 0.55 |
| Sleep disturbances | 1.41 (0.60) | 1.38 (0.57) | 2.25 | **0.02** |
| Sleep medications | 0.29 (0.80) | 0.33 (0.86) | -1.90 | **0.049** |
| Daytime dysfunction | 0.85 (0.71) | 0.92 (0.70) | -4.14 | **< 0.001** |
| ISI score | 8.34 (5.45) | 7.73 (5.39) | 5.49 | **< 0.001** |
| MEQr score | 15.31 (3.66) | 15.44 (3.68) | -2.53 | **0.01** |
| *Psychological status* | Mean (SD) |  | t | P |
| BDI-II score | 12.46 (8.96) | 12.35 (9.41) | 0.57 | 0.57 |
| PSS-10 score | 17.99 (7.41) | 18.70 (3.74) | -4.17 | **< 0.001** |
| STAI-X1 score | 48.58 (9.17) | 46.78 (9.39) | 8.24 | **< 0.001** |
| *Sleep/Psychological disturbances* | N (%) | | $\chi^2$ | P |
| Poor sleep | 1116 (60.16) | 1110 (59.84) | 0.04 | 0.84 |
| Moderate/severe insomnia | 270 (13.41) | 226 (11.23) | 6.56 | **0.01** |
| Severe depression | 352 (19.06) | 370 (20.03) | 0.90 | 0.34 |

## Discussion

In line with the literature about the first wave (e.g., Salfi et al.,2020b), we confirmed a higher predisposition of female gender and low education level to develop sleep disturbances during our second wave assessment. The healthcare workers confirmed their vulnerability to insomnia symptomatology during the second wave (Pappa et al., 2020), while the elderly confirmed higher predisposition to poor sleep quality (Mander et al., 2017).

The circadian preference turned out as a crucial predictor of the sleep outcome, consistent with pre-pandemic literature (Adan et al., 2016): evening-type individuals showed a higher predisposition to poor sleep quality and moderate/severe insomnia symptoms, while morning chronotype emerged as a protective factor. The follow-up assessment of our investigation took place during a period of lighter restraining measures. In Italy, during the second wave of contagion, a regional lockdown was adopted, and we failed to highlight any difference according to the rigidity of the restraining measures adopted. This result pointed to a detrimental effect of the pandemic period itself, regardless of the restrictions in force. We demonstrated a higher risk for insomnia of individuals who lived with children. School activities in presence were suspended in Italy during the follow-up measurement while working activities partially continued (especially in orange and yellow zones). This may have created a difficult situation to manage for parents, explaining our results.

Being at-risk subjects for COVID-19 infection was associated with a higher predisposition to poor sleep quality and moderate/severe insomnia symptoms, while living with high-risk people for COVID-19 predicted a higher probability of experiencing poor sleep quality. These results could reflect a higher fear of contagion and worries experienced by these individuals, which triggered sleep disturbances. Having contracted the COVID-19 or having a relative/close friend infected before the prior two weeks of the survey participation constituted a risk factor for insomnia and poor sleep quality, respectively, pointing to a long-term impact of these events. Notably, optimism for the future due to the vaccine’s arrival emerged as a protective factor against the insomnia exacerbation. Therefore, it seems that the vaccination campaign prospective could be itself beneficial for sleep health.

The pandemic has been going on for months, and the healthcare emergency was complemented by unprecedented economic and social crises. In this context, more than one-third of the sample reported a negative economic impact of the current situation, and six of ten of the respondents experienced a consistent impairment of their social relationships. Both these outcomes turned out to be risk factors for the exacerbation of sleep disturbances. Finally, the smartphone overuse before the sleep onset emerged as a risk factor for sleep disturbances. This finding is putatively ascribable to the well-known detrimental effect of backlit screen exposure before sleep time on the circadian system, as well as to the alerting effects of digital engagement. Notably, the increased evening usage of electronic devices has been already proposed as a mediator of sleep deterioration during the March–April lockdown (Salfi et al., 2020a).

Comparisons between data from the two outbreak waves displayed an articulated framework.

We showed improved insomnia symptoms, reduced prevalence of moderate/severe insomnia conditions, and reduced anxiety. However, the present investigation confirmed the alarming situation highlighted during the first wave of COVID-19 (Jahrami et al., 2020, Salfi et al., 2020b), since the majority of the sample consisted of poor sleepers and this prevalence (∼60%) remained stable between the two pandemic waves.

Despite the invariance of sleep quality between the two assessments, we showed several differences as concerns the PSQI sub-components. Subjective sleep quality, sleep latency, and sleep disturbance dimensions improved. However, the improvements were compensated by reduced sleep duration, more severe daytime dysfunction and higher sleep medication use. Moreover, the participants went to bed more than half an hour earlier and woke-up almost an hour earlier than during the March–April lockdown. These results were accompanied by a significant shift towards the morning chronotype. In light of these results, it should be acknowledged that the first lockdown period was characterized by a substantial reduction of the social jetlag due to weaker social and working obligations (Korman et al., 2020). Our findings suggest that the social jetlag returned to negatively influence Italians’ sleep since the second assessment period was marked by a substantial resumption of daily working and activity routine.

Finally, we confirmed the severity of depressive symptomatology and the alarming prevalence of severe depression conditions (∼20%) of the March–April lockdown. Remarkably, all these results were obtained in a context of increased perceived stress, putatively ascribable to the prolonged emergency period.

In conclusion, our study demonstrated that the impact of the COVID-19 pandemic persists on both sleep and mental health, although the second wave of contagion has been faced using lighter restraining measures. Therefore, vigilance is still required, and large-scale interventions should be performed to counteract the chronicity and exacerbation of sleep and psychological disturbances, especially for the categories identified as at-risk in the present study.

## Data Availability

The data underlying this article will be shared on reasonable request to the corresponding author.

## Funding

This research did not receive any specific grant from funding agencies in the public, commercial, or not-for-profit sectors.

## Author contributions

**Federico Salfi:** Conceptualization, Methodology, Investigation, Data curation, Formal analysis, Writing - original draft, Writing - review & editing. **Aurora D’Atri:** Writing - original draft, Writing - review & editing. **Daniela Tempesta:** Writing - review & editing. **Michele Ferrara:** Conceptualization, Methodology, Investigation, Supervision, Writing - review & editing.

## Declaration of competing interest

None of the authors have potential conflicts of interest to be disclosed. All authors have seen and approved the manuscript.

## Notes

### Competing Interest Statement

The authors have declared no competing interest.

### Author Declarations

The study has been approved by the Internal Review Board of the University of L Aquila (protocol n. 43066).

